# Training-of-trainers program for community health workers involved in an innovative and community-based intervention among goldminers in the Guiana Shield: a quality and effectiveness evaluation

**DOI:** 10.1101/2023.10.10.23296813

**Authors:** Carboni Carlotta, Jimeno Maroto Irene, Galindo Muriel, Plessis Lorraine, Lambert Yann, Bardon Teddy, Vreden Stephen, Suárez-Mutis Martha, Miller Bordalo Jane, Douine Maylis, Sanna Alice

## Abstract

**Introduction:** An innovative and community-based intervention is implemented in the Guiana Shield to eliminate malaria among people involved in artisanal and small-scale gold mining. The intervention is possible in the field thanks to community health workers (CHWs) who are previously trained to master all intervention’s procedures, including training gold miners to treat themselves for malaria. This article describes thus a training-of-trainers (ToT) program for CHWs, as well as the method and results of its evaluation in terms of quality and effectiveness. **Methods**: A mixed-method case study was implemented in two steps and based on knowledge survey, satisfaction test, observations and semi-structured interviews. Quantitative and qualitative data were triangulated. **Results**: The project team’s previous experience and the Guiana Shield countries’ commitment to the WHO E-2025 initiative were identified as levers for the quality of the ToT training, while the complexity of the project context was a challenge. Group dynamics and adaptations were found to be central elements of a high-quality training program. CHWs’ satisfaction was elevated especially regarding training format and learning results. Improvements on knowledge level demonstrated good effectiveness of the training. Nevertheless, some difficulties persisted regarding certain tasks to be carried out by CHWs during the intervention procedure. Further on-the-job training permitted to address them, improving CHWs’ practices in the field. **Discussion**: High-quality, effective, and appropriate training programs are required for effective and sustainable intervention involving CHWs profiles. Training design is a crucial point to attend quality and effectiveness. ToT model has been shown to allow a high level of satisfaction, good learning results and satisfactory implementation in the field. Initial and continuing training is an indispensable continuum to sustain good practices in the field and CHWs’ motivation. Training evaluation permits to standardize methods and facilitate transferability of successful efforts.

## 1 Introduction

Although since 2000 the malaria burden considerably decreased in the Americas, it is still endemic in the Guiana Shield (1–3). In this area, the major malaria reservoir is represented by a mobile, and hard-to-reach population challenging the fight against malaria (4,5). This population is mainly represented by Brazilians (95-98%) involved in artisanal and small-scale gold mining (ASGM) in the Amazonian rainforest, where they live in a situation of vulnerability with reduced access to the health care system (6,7).

Despite the complexity of reaching populations suffering from social inequality, strategies have been developed to provide health care access (8). Indeed, at a global scale community-based interventions play a fundamental role in improving population health, and increasing access to health care and services (9,10). Worldwide, a range of effective interventions involving community health workers (CHWs) exists, especially with the aim of fighting malaria (11,12). In malaria elimination programs, CHWs are often involved in case management, malaria surveillance, diagnosis, treatment, prevention and health promotion activities (13).

A complex, innovative and community-based intervention has been implemented in 2018-2020 as a result of a scientific partnership established between Brazil, French Guiana and Suriname focusing on populations involved in ASGM in the Guiana Shield in situations of extreme isolation from health services (14). The aim was the evaluation of a strategy that consisted in providing access to diagnosis and treatment through the distribution of kits (called “malakits”), associated with a training to use it correctly (15). The malakit was composed of self-testing kits with rapid diagnostic tests (RDTs) and self-treatment kits in case of malaria This strategy was evaluated as effective and acceptable by the community, and has been scaled up in Suriname (16–18).

Started in 2023, another intervention’s evaluation study – Curema – expands the foundations of the Malakit project to also include the radical cure of *Plasmodium vivax* (19,20) . Curema intervention consists of two modules offered at inclusion sites located in Brazil and Suriname, in resting and logistical places across the border in French Guiana, regularly frequented by people involved in ASGM in French Guiana: the “malakit” module and the “radical cure” module. The latter aims to prevent relapses and thus reduce the number of human hosts capable of transmitting the parasite; asymptomatic individuals at risk of carrying *P.vivax* hypnozoites are offered presumptive treatment with chloroquine and aminoquinolines.

The intervention is made possible by CHWs, specifically recruited for the Curema project, and working in pairs on the field at the inclusion sites. As part of the Curema project, CHWs are responsible for enrolling participants and providing health education to participants followed by individual training during the inclusion process (**Figure 1**). Furthermore, they carry out collective outreach activities on malaria in the community.

**Figure 1:**
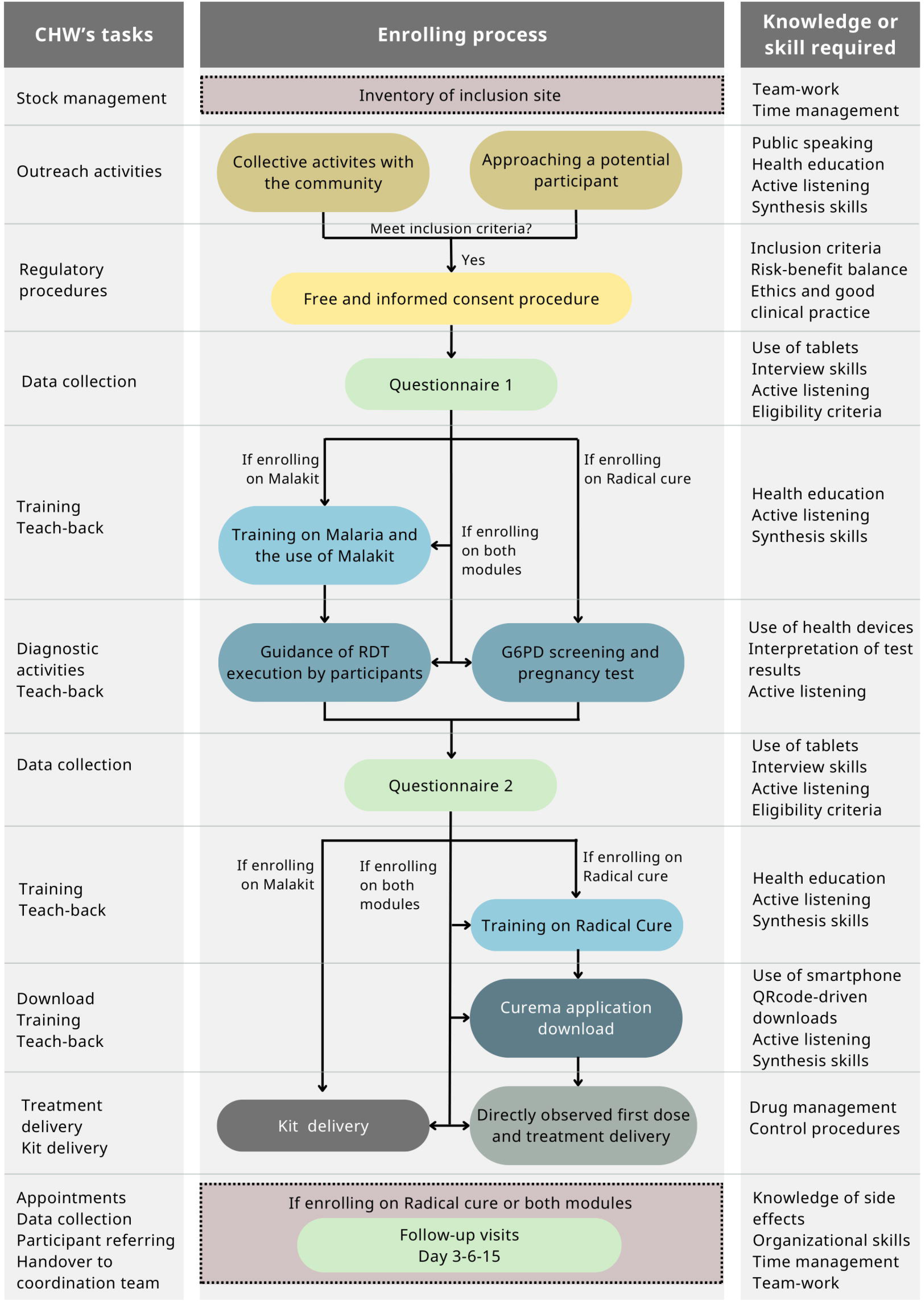
CHWs tasks over Curema enrolling process and knowledge/skills associated with.

Before the launch of the project, the CHWs participated in a training program which enabled them to take part in a bidirectional working space, to master health research procedures, to present information effectively, to lead activities that reinforce participants’ learning, and to collaborate on the design of working tools. Moreover, they were supported during the gradual launch of the project in the field and regularly supervised during the whole project implementation.

Despite the large integration of CHWs in public health interventions, little is known about best practices for training them. To ensure effective CHWs activities, it seems vital to provide a good training and a continuous professional development through supervisions. Nevertheless, literature is often lacking evidence on the implementation and evaluation of training programs. A number of theoretical frameworks have been developed for evaluating this process, but the introduction of standardized evaluation methods in the context of training evaluations is increasingly necessary (21– 24).

Therefore, it seems crucial to share findings and experiences in this domain with the scientific community, in order to facilitate the transferability of successful efforts.

This present article aims to illustrate the results of an evaluation of the quality and effectiveness of the training program offered to CHWs involved in the Curema project.

## 2 Materials and methods

### 2.1 Description of the Curema training program

#### 2.1.1 CHWs’ recruitment

The recruitment process was conducted separately by local partners for Brazilian and Surinamese inclusion sites with the support of the project’s promotor, the Hospital of Cayenne, French Guiana. Their profile is similar to CHWs involved in malaria intervention in other countries (25,26). CHWs’ recruitment criteria were: being over 18 years of age, belonging to the community involved in ASGM or knowing it closely, speaking fluent Portuguese, being able to use information technology tools (tablets, smartphones, etc.), living in or being prepared to move to one of the project locations for inclusion sites. Moreover, a technical diploma as nurse-assistant was an asset for CHWs assigned to Brazil.

The CHWs working at the Brazilian inclusion sites were employed by the NGO DPAC-Fronteira (*Desenvolvimento, Prevencao, Acompanhamento e Cooperacao de Fronteiras*; Development, Prevention, Support and Cooperation on Borders) and were only involved in activities linked to the Curema project. DPAC-Fronteira selected CHWs before they underwent the training program. At the Surinamese inclusion sites, the CHWs were employed by the Surinamese research foundation SWOS for the Curema project (half-time) and by the Surinamese National Malaria Elimination Programme (NMEP) for the second half-time as “malaria service deliverer” (MSD). This MSD role included in malaria case management (diagnosis, treatment, notification, and investigation) as well as in distributing mosquito nets, in remote areas related to ASGM (27). CHWs for Suriname inclusions sites underwent a two-steps recruitment process, consequently a larger number of potential CHWs was invited to attend the training program. Most CHWs invited to attend the Curema training program, already functioned as MSD in the mining fields. It was made clear to all trainees, that participating in the training was no guarantee for being selected to work in the Curema project. Final CHWs were selected after the end of the initial training, just before the launch of the project in the field.

Prior to the Curema training program, CHWs undertook basic training sessions separately as a pre-work. In Brazil, a basic one-day training on malaria was provided by Fiocruz and a training on NGO work and community mediation was provided by DPAC. In Suriname, CHWs received initial and refresher training on malaria prevention, diagnosis, and treatment techniques by NMEP.

#### 2.1.2 Design and development of the training program

With the aim of making CHWs able to master all the procedures and tasks they need to carry out, the Curema training program was designed using a training-of-trainers (ToT) model with two components: an initial training and an on-the-job training (**Figure 2**).

**Figure 2:**
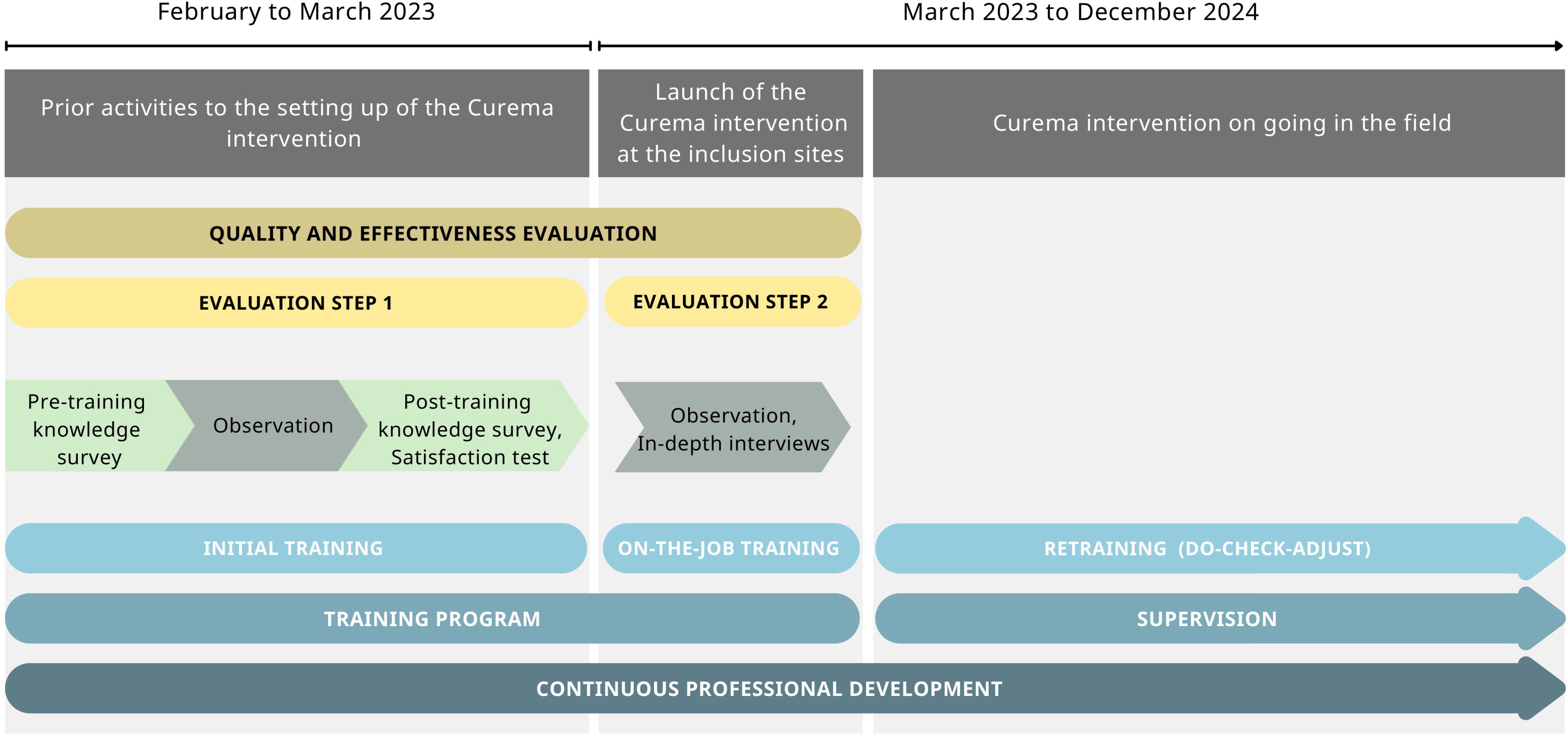
Timeline and flowchart of the training design and training evaluation over Curema intervention phases.

Adult learning principles were applied to design the training program focusing on practice-based activities to be appropriate for CHWs’ needs and to give them the opportunity to practice (28). To be more effective, and to reach all of the audience, varied activities and methods were implemented mobilizing cognitive, affective, psychomotor learning principles, and the visual, auditory and kinesthetic (VAK) learning model (29–31). Moreover, the training program was tailored and centered on CHWs. Therefore, the training program was conducted in Portuguese using learning tools targeting on an audience with a low level of scholarity.

All partners and organizations involved in the Curema project participated in the training program, increasing contributions from all staff and from professionals with different experiences and expertise. Trainers used a horizontal approach to make CHWs feel respected and to make the learning environment safe and supportive. To be more responsive to the needs of the CHWs and the community in the field, a bidirectional approach was used, and CHWs were asked to collaborate in the design of operating tools.

The initial training component took place in Paramaribo, Suriname, jointly by the entire team of CHWs both from Suriname and Brazil. Considering the type and number of topics to be addressed, the initial training was developed for three-weeks to allow enough time to ensure an effective knowledge transfer, an exhaustive practice of skills and new procedures, an appropriate adaptation of tools, and a good team and capacity building.

The on-the-job training component took place at each inclusion site and lasted two to three days. Its aim was to support CHWs while the project was gradually rolled out in the field and the first inclusions carried out.

The components included on the ToT program, as Centers for Disease Control and Prevention recommend, are shown in **Table 1** (32).

**Table 1:** The ToT components included on the Curema training program.

| ToT component | Pre-assessment | Pre-work | Training agenda | Facilitation manual | Modelling of skills and tasks |
| --- | --- | --- | --- | --- | --- |
| **Aim** | Identifying pre-training knowledge and skills of the CHW, to determine training needs | Providing CHWs with the knowledge and background needed before the training program | Providing some degree of control over what, when and how CHWs will learn during the training program. | Organizing tools and improving the capacity of CHWs to transfer knowledge | Enabling CHWs to form a clear idea of how to carry out a task by showing them how to perform a skill while describing each step |
| **Initial Training** | Pre-training knowledge survey | Topics: basic knowledge about malaria (diagnosis and treatment), epidemiology in the Guiana Shield and malaria control strategies<br>Pre-work training sessions took place separately for the CHWs working on the inclusion sites located in Brazil or Suriname, due to different recruitment frameworks. Those sessions were provided respectively by two different organizations, due to legitimacy of respective countries | Provided on the first day of the training | Audio-visual tools (i.e. a step-by-step tutorial about how to perform a Malakit RDT) were uploaded to tablets<br>Visual sheets for explanation support were put together in a CHW binder<br>Games and teach-back cards were also provided as teaching tools | Following training units:<br>Audio-visual tutorial for Malakit RDT and for G6PD test;<br>Information Education and Communication (IEC) strategies and tools use;<br>Inclusion process simulation role-play;<br>Informative note and informed consent procedure role-play |
| **On-the-job training** | Post-initial-training knowledge survey<br>Feedback of the trainers regarding the initial training program outcomes | Initial training | Not applicable | The same tools and CHW blinder with the addition of some updated sheets | Not applicable |

| ToT component | Adults learning principles | Skill practice | Feedback | Action planning | Planned follow-up support |
| --- | --- | --- | --- | --- | --- |
| **Aim** | Diversifying activities based on VAK Model and Bloom's Taxonomy, enhancing experience, designing a problem-centered training program to reach CHWs and make the training effective | Providing the opportunity for the practice of selected training activities or contents. | Collecting CHWs' feedback regarding the learning process to be adjusted. | Taking CHWs through the process of creating or adjusting tools and practices for the intervention to succeed. | Strengthening the knowledge and skills of CHWs, so they can be applied effectively. |
| **Initial Training** | Implementation of games, practical simulations, video making, and a variety of activities with movements needed<br>Activities in small working groups composed of heterogeneous profiles for skills and personalities | Practice of:<br>data collection, tablet use, G6PD test execution, Malakit RDT execution monitoring, participants and community training, health education, public speaking, inclusion process simulations, installation of Curema application for smartphone | Daily debriefing at the beginning of the day regarding previous content, doubts, difficulties.<br>Use of a checklist to help on completing the whole inclusion process<br>Satisfaction assessment and discussion group on the last day | Information Note video - for the signature of the ICF (Informed Consent Form)<br>Feedback about Questionnaire | Continuous professional development through on-going training for the intervention launch on the inclusion sites. |
| **On-the-job training** | Directly linked to on-site work and problem-solving | Inclusion process, community outreach activities | Debriefing together at the end of inclusion process (either real process or simulations) | CHWs' initiatives encouraged (group of discussions, projection of promotional videos, marauding and outreach activities, etc) | Continuous professional development and targeted follow-up:<br>(i) regular supervision visits<br>(ii) remote support via a specific WhatsApp group |

### 2.2 Evaluation of the quality and effectiveness of the training program

#### 2.2.1 Framework

Evaluation was carried out for the quality and effectiveness of training.

Quality was defined by the International Standard Vocabulary as “all the characteristics of an entity that influence its ability to satisfy expressed and implicit needs” (33). In this context, authors referred to “needs” as the outcomes to be achieved by CHWs through the training program.

Effectiveness corresponded to the extent to which the training outcomes are achieved. According to Kirkpatrick’s theoretical framework, effectiveness is assessed at 4 levels: reaction, learning, behavior, and system (34).

The elements to be evaluated were defined as follow (24):

- Quality assessment

`o` Context: external factors which can influence the training program, both as obstacles and levers and which cannot be controlled by the training deliverers.
`o` Input: internal conditions under which the training program takes place (eg. methods used by trainers, CHWs’ prior knowledge, group dynamism, etc.).
`o` Process: the training delivery process, including the adaptation, dose delivered, CHWs reached.
- Effectiveness assessment

`o` Reaction: level of CHWs’ satisfaction in terms of dose and content.
`o` Learning: changes in level of knowledge, know-how and interpersonal skills.
`o` Behavior: level of concordance of CHWs’ practice with the reference (Curema protocol)
`o` System: impact of CHWs’ practice on community health education

#### 2.2.2 Methods

The design of the training evaluation consisted of a mixed-method case study, executed in two steps as shown in **Figure 2**. Firstly, during the initial training in Paramaribo developed in a classroom environment, and secondly during the on-the-job training during the launch of the intervention on inclusion sites. First step is nurture by quantitative and qualitative methods, while second step correspond to qualitative method. The quantitative component provided computable data on satisfaction, knowledge improvement and practice, while the qualitative component enabled giving context and an in-depth explanation of the quantitative results.

Data were collected through knowledge surveys, a satisfaction test, observations, and semi-structured interviews, and analyzed as follows (**Table 2**):

The analysis was conducted using a theoretical framework conceptualized for the evaluation of training programs for the control of infectious diseases in cross-border contexts, by the Centre for Infectious Disease Control, National Institute for Public Health of Health of The Netherlands and The EU Healthy Gateways Joint Action (24).

**Table 2:**
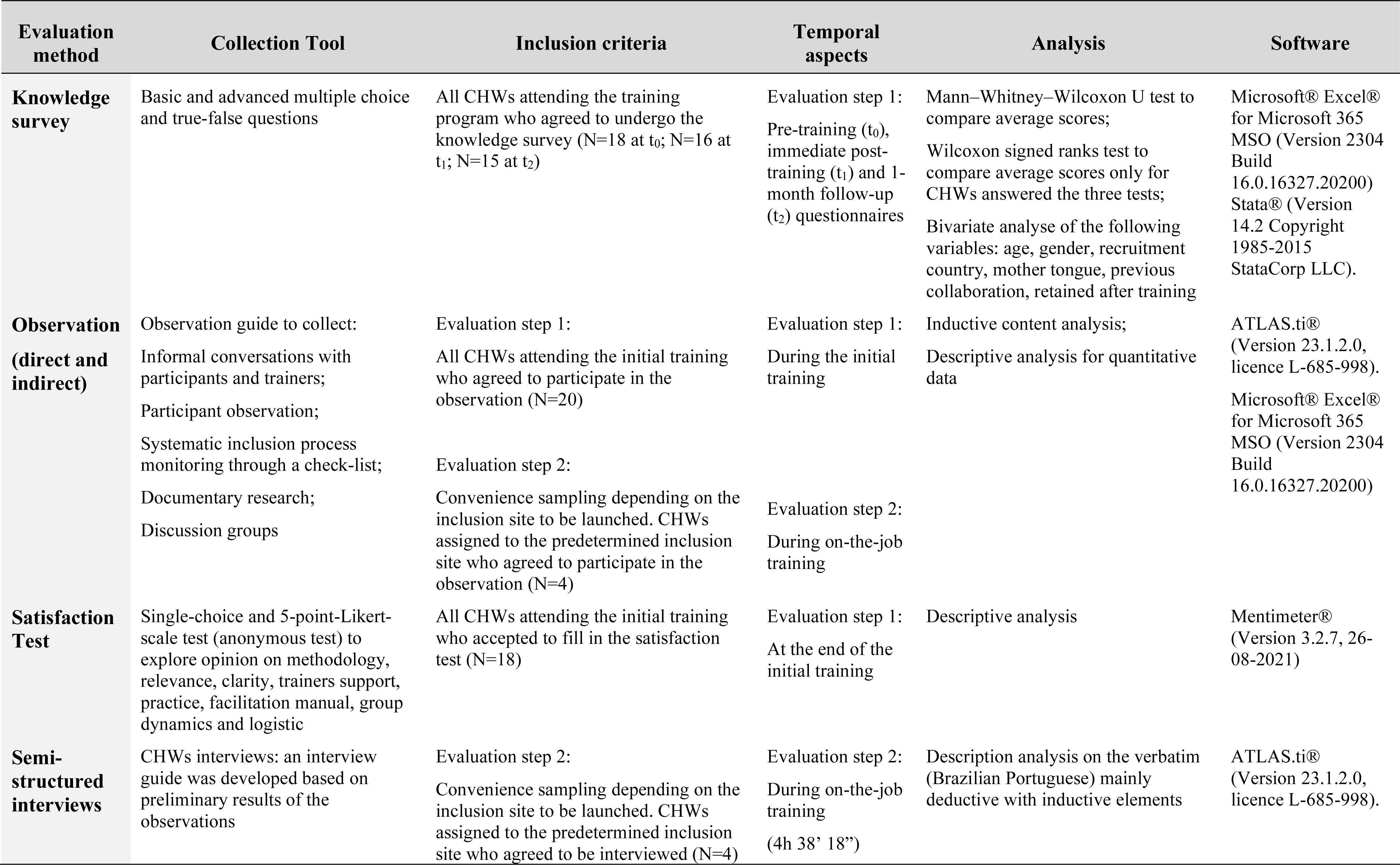
Methodology used to evaluate quality and effectiveness of the Curema training program.

#### 2.2.3 Ethics

Curema protocol was approved by the ethics committees of the countries involved in the project (Brazil: CONEP:5,507,241 and Suriname: CMWO05/22). The evaluation of quality and effectiveness of CHWs’ training program is part of the secondary objectives of the Curema protocol.

According to the article R1121-1 of the French Public Health Code, the regulatory classification of this evaluation is a “research not involving the human being” (outside the scope of the *Loi Jardé* or *RNIPH*) because it “aims to evaluate the methods of practice of healthcare health professionals or teaching practices in the field of health”(35). A collective oral information was reached out and participants’ non-opposition was collected. For semi-structured interviews, consent was requested.

The evaluation was conducted in accordance with the principles of the Declaration of Helsinki. Data collected were recorded in accordance with French law and the European Union General Data Protection Regulation. Data collected, field notes and transcribed recordings were anonymized and stored on a secure server.

## 3 Results

The initial training took place between February the 13^th^ 2023 and March the 03^rd^ 2023. On-the-job training stared with the launch of the intervention at the first inclusion site on March the 13^th^ 2023 and lasted two to three days per site. The evaluation was carried out between February the 13^th^ 2023 and April the 03^rd^ 2023.

### 3.1 CHWs profile

A total of 20 CHWs were invited for the training program, 14 from Suriname employer-organizations and 6 from Brazil employer-organization. The characteristics of the trainees are summarized in **Table 3**. The majority were women over 40, with Portuguese as their mother tongue. Five CHWs had Spanish as their mother tongue, but could proficiently speak and understand Portuguese. Less than a quarter had previously worked in research. CHWs profiles were different in Brazil and in Suriname because of different recruitment criteria. Of the 20 CHWs, 4 took part in individual semi-structured interviews during Evaluation Step 2.

**Table 3:** Characteristics of CHWs who attended initial training program in Paramaribo and who participated to semi-structured interviews.

| CHWs characteristics |  | Involved in the initial training in<br>Paramaribo<br>(N=20) |  | Involved in semi-structured<br>interviews<br>(N=4) |  |
| --- | --- | --- | --- | --- | --- |
|  |  | Percentage<br>(%) | Number<br>(n) | Percentage<br>(%) | Number<br>(n) |
| Sex | Male | 20.0 | 4 | 25.0 | 1 |
| Age (years) | 18-40 | 20.0 | 4 | 25.0 | 1 |
|  | >40 | 80.0 | 16 | 75.0 | 3 |
| Country of the<br>employer-organization | Brazil | 30.0 | 6 | 0.0 | 0 |
|  | Suriname | 70.0 | 14 | 100.0 | 4 |
| Mother tongue | Portuguese | 75.0 | 15 | 100.0 | 4 |
|  | Spanish | 25.0 | 5 | 0.0 | 0 |
| Previous collaboration with<br>the employer-organization | Yes | 50.0 | 10 | 100.0 | 4 |
| Previous work experience<br>in research | Yes | 20.0 | 4 | 0.0 | 0 |
| Belonging to ASCM- related<br>community | Yes | 70.0 | 14 | 100.0 | 4 |
| Retained after training | Yes | 75.0 | 15 | 100.0 | 4 |

### 3.2 Results of the quality evaluation

#### 3.2.1 Contextual levers and difficulties

Two elements were identified as levers for the training. The first is the political, economic and intellectual environment in which the Curema project – and consequently the training program – navigates. In fact, Suriname and French Guiana have committed to eliminate malaria from their territories by 2025 (World Health Organization’s E-2025 initiative) and Brazil by 2035 (1,36,37). There is no doubt that this commitment ensured good conditions for the start of Curema intervention and its training program. Second, previous lessons learned during Malakit project by our team in training the same community in a similar context and partners experiences were precious to guide the design and development of the ToT program, particularly regarding the methodology, the time, the appropriate tools for the audience (38).

On the other hand, the complexity of Curema project also influenced the training because of a large number of interacting components that are sometimes difficult to align in order to make possible the training program development (39). In fact, Curema is an international project which means several organizational levels, country-specific features for implementation on the field, communication in different foreign languages depending on the actors, logistical issues linked to differences in bureaucracy and regulatory requirements between countries (40). Moreover, it was important to consider that the number and level of articulations of the activities required by CHWs complexified the training program design. Finally, differences on recruitment and employment frameworks made CHWs group composition heterogeneous: previous baselines knowledge were dissimilar due to previous experiences and to a pre-work organized separately for Brazil and Suriname.

#### 3.2.2 The importance of group dynamics

Among the CHWs who attended the training program, 4 (20.0%) had already worked in the research field and 10 (50%) had already collaborated with one of the three employer-organizations involved in the Curema project. We observed that, for these CHWs, prior knowledge of certain procedures increased mastery of the subject and reduced the time needed to learn. In addition, thanks to a participatory approach to training and the inclusion of practical sessions of pair and group work in the schedule, peer education dynamics took place. The attention in the choice of the classroom and the setting is also decisive in the generation of these dynamics. Thus, these CHWs became a resource for supporting training. However, certain disadvantages were also identified such as the transfer of some erroneous automatisms already adopted in their practice. Feedback activities allowed to readjust practices.

The observed interpersonal relationships among CHWs had a strong influence on group dynamics for learning, active participation in training and engagement in the project. Because of the different recruitment framework, at the beginning of the training program CHWs appeared to form two distinct groups. Based on team building and adult-learning principles, more collaborative activities were implemented in order to facilitate exchanges and integration. Some spontaneous team-building initiatives were also pointed out.

> “Afterwards, the group work was done, which […] allowed people to feel more in tune with each other and communication improved […] which was fun and at the same time a joint learning process, it was very good." (Interview verbatim, CHW from Suriname)

#### 3.2.3 A flexible and evolutive process

The training was delivered in 105 hours over a period of three consecutive weeks (**Table 1**). The training program was designed to be part of a continuous professional development approach centered on CHWs. Thus, it was flexible and evolutive. Numerous adaptations were made to ensure constant improvement of CHW knowledge and practice and to perfect training (schedule, activities, tools) and project elements. Indeed, many of those elements had already been co-designed through community involvement. The bidirectional working space during the training sessions enabled to adapt vocabulary and content of the inclusion questionnaire, to add supplementary material in the facilitation manual, to make a video for the informed consent procedure. As they reported during informal conversations and semi-structured interviews, CHWs greatly appreciated the opportunity to be part of this evolution process.

### 3.3 Results of the effectiveness evaluation

#### 3.3.1 Reaction: participant’s satisfaction

Overall, CHWs affirmed to be very satisfied with the training. The methodology used for the training was particularly appreciated by CHWs with reference to activities using kinesthetic learning model, practice-based activities, group-work and bidirectional approach. All items (**Table 2**) explored on the scale of 1-5 ranging from "not at all satisfied" to "completely satisfied", were ranked over 4. Satisfaction is shown to be directly linked to learning:

> “It was fun, creative, and useful for memorizing, because you remember [*what you learn using these techniques*], isn’t it? We don’t forget. The games were very good" (Interview verbatim, CHW from Suriname).

Indeed, the 61.1% of the 18 CHWs answering the satisfaction test declared had reached "optimum mastery", the 27.8% reported "sufficient mastery but needed support during the launch in the field"; and only one person felt "insufficient mastery" asking for "more time for training".

CHWs were observed to be satisfied also during on-the-job training. In fact, some of them stated trainees indicated that the sessions provided the knowledge and practice needed to start the job in the field.

#### 3.3.2 Learning: evaluation of knowledge improvement

The evaluation found a knowledge improvement with a significant increase in the average scores of the test between pre and post survey with a p<0.05 both for test scores at t_0_-t_1_ and t_0_-t_2_ (**Figure 3**). On the contrary, differences in scores between tests at t_1_ vs t_2_ were not significant (p=0.26). The same trend was shown for analyzing individual scores’ increase for CHWs which underwent the knowledge survey at t_0_, t_1_, and t_2_. The bivariate analysis for age, sex, recruitment country, mother tongue and previous collaboration did not show any differences in these categories, apart from a better basic knowledge in the t_2_ test among those with previous experience in research field (p<0.05).

**Figure 3:**
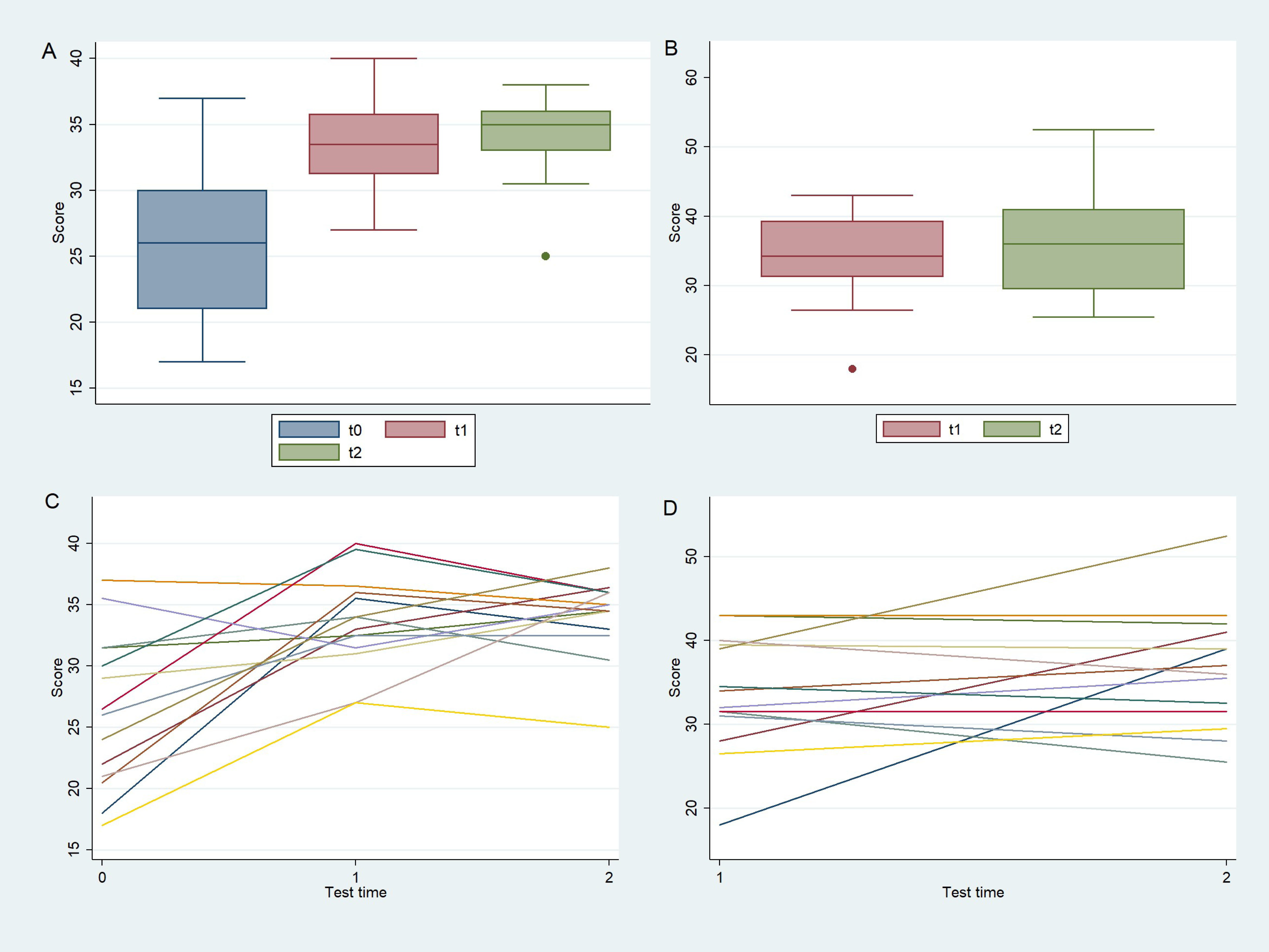
Evolution over time in the knowledge survey score for basic knowledge (at t_0_, t_1_, and t_2_) among all CHWs (**A**), for advanced knowledge (at t_1_, and t_2_) among all CHWs (**B**), for basic knowledge (at t_0_, t_1_, and t_2_) per CHWs (**C**), for advanced knowledge (at t_1_, and t_2_) per CHWs (**D**).

Systematic inclusion process monitoring through the checklist pointed out that two-thirds of observed inclusion simulations (n=16) contained inaccuracies in the step-by-step process. Nevertheless, inaccuracies often concerned the same steps. The highest rate of inaccuracy was found for the explanation of the patient information note for informed consent procedure that was inaccurate in 29.2% of the observed inclusion simulations, followed by the process of installing the Curema smartphone application, which had an inaccuracy rate of 16.7%. These results are in line with the results of the qualitative analyses based on informal conversations and interviews with CHWs who also reported reasons why those two steps were difficult in their opinions. First, the information note, using scientific terms and constructions, was dense of complex concepts and difficult to be understand for people with poor level of scholarity.

> “What really frightened us, I can say for myself and for several other people, was the informative note […] What really complicates things for me is the informative note.” (Interview verbatim, CHW from Suriname)

Second, the Curema smartphone application was not enough explored during the training program. Indeed, at that time, some technical developments of the application were in progress. Thus, the final version and procedure to use it were not integrally exposed.

#### 3.3.3 Behavior: from the training to the field

During the observed launch at the inclusion sites, the level of concordance of CHWs’ practices with the Curema standard procedures was adequate even if, during on-the-job training, further support by trainers was needed to guide CHWs towards autonomy. Moreover, CHWs expressed the need for more time and fieldwork to master the step-by-step process and re-elaborate concepts for an effective knowledge transfer to the community.

Constructive professional relationships, good team working, and a sense of belonging to a common cause were also pointed out by CHWs as factors able to influence behavior in the field, leading to engagement and proactive participation in the intervention.

> “If you don’t feel good in the team, this can have an influence on your motivation, for example, in the way the work is run.” (Interview verbatim, CHW from Suriname)

## 4 Discussion

High-quality, effective, and appropriate training are required for effective and sustainable intervention involving CHWs profiles. This involves a great deal of effort in development, implementation, and evaluation. Moreover, this study highlights the importance of quality to achieve effectiveness of a training program. ToT model has been shown to allow a high level of satisfaction, good learning results and satisfactory implementation in the field.

### 4.1 Limits of our study

The evaluation was carried out by researchers previously involved in the design, development, and implementation of the training. Despite this could lead to attribution bias, several data sources – both quantitative and qualitative – were used for the evaluation, enabling data triangulation and increasing internal validity.

The Kirkpatrick’s theoretical framework used recommends to assess effectiveness at 4 levels: reaction, learning, behavior, and system. The higher the impact in terms of effectiveness, the better the achieved the final training outcome (24). Nevertheless, the assessment at the system level is more challenging and we were unable to evaluate it. A further evaluation of learning transfer to the community at the end of the intervention could be realized by the post-intervention cross-sectional survey and would be very useful.

### 4.2 Training design: a crucial step

Developing a high-quality training program demands careful considerations of the learning methods and modules to be designed. We designed the training program on a ToT model based on adult-learning principles. Consequently, the training program contained ample interactive sessions including group discussions, role plays, practical activities, and simulations. This methodology is considered more effective in comparison to traditional teaching approaches, especially among CHWs with poor level of scholarity (9,41). Despite complexities, we can affirm that the quality of the training was good and met the needs for the implementation of the Curema intervention in the field. The significant increase in pre-post scores obtained at the knowledge survey suggests the achievement of theorical concepts. Moreover, the ample panel of activities helped increase the satisfaction level, in accordance with the literature (24).

A crucial point to be discussed is adaptations. During the training program, several adaptations were necessary to be adopted both for training program and for the intervention on the field. We could even say that one of the aims of the training was to enable these adaptations to be made, thanks to the bidirectional space set up with the CHWs. Despite the balance between adaptation and fidelity in implementation science is still a topic of debate (42–44), several examples in literature described helpful adaptations: Miller et al. adapted terminology of their intervention to reduce the possibility of misinterpretation of their study population, while Holtrop et al. shown training program or recruitment adaptations due to outside forces or made to improve feasibility or engagement (45,46). A review by Stirman et al. identified most common reasons intervention adaptation were to address language, cultural differences, literacy, or situational constraints (47). In our experience, we can argue that adaptive aptitude, embedded in a community-based approach, may have enabled our intervention to be more appropriate, effective, and sustainable by fostering the bidirectional transfer of knowledge, the empowerment of CHWs as a central actor of the intervention, and their engagement in the Curema project.

To be appropriate, trainings of the CHWs should take place in their work environment to practice in a real-life situation (48,49). Our quantitative and qualitative results suggested that, although many simulations were carried out during the initial training, real practice in the field during on-the-job training permitted CHWs to improve mastery of procedures and to appropriately perform participants’ inclusions and education activities regarding malaria. Similar results have been shown in previous experiences among the same community and intervention patterns by Galindo et al. who concluded that practical training in the field was the most appropriate learning method for CHWs (38). Besides, a scoping review describing training, supervision and quality of care in community-based programs in different contexts from the studied one, also concluded that training in the field and on-site have been shown to improve CHWs’ practices (50).

Another important element to address is team-building, which has been found out as a crucial element for learning process and daily work on the field. During the training, the team-building activities helped to motivate CHWS and encouraged peer learning. Moreover, the sense of unity jointly with the engagement towards the goal of eliminating malaria increased CHW’s motivation in the daily working context, particularly in regions where malaria is in the process of being eliminated. Similar results showing how relationships with other cadres and actors could play a major role on sustain motivation are reported in literature (51,52).

### 4.3 Evaluation

Nowadays, limited number of studies exists regarding quality and effectiveness evaluation of training programs, especially those regarding infectious diseases in cross-border areas and those addressed to CHWs. Indeed, a recent review of the literature analyzed 62 articles on the evaluation of ToT, and found only 5 in the field of infectious diseases and in cross-border contexts, with heterogeneous and non-standardized methodologies (24). Reviewing articles on CHWs-based programs for cardiovascular disease management in low-income and middle-income countries with the aim of evaluate training program effectiveness, Abdel-All and al. found only 8 articles reporting training details on evaluation, among 90 eligible for their systematic review (53).

Scientists call for studies examining the effectiveness and characteristics of training programs for CHWs to provide reliable evidence, to avoid replication of programming difficult, to ensure high-quality of training programs, to make possible standardization on the evaluation (53–57). This study shows as developing methodology for training evaluation is possible, feasible and necessary to ensure advancements on this field. Evaluation of training programs must be integrated in all CHWs-based health intervention.

### 4.4 Initial and continuing training: an indispensable continuum

ToT is a good predictor of the long-term sustainability of public health initiatives (23). This ToT program was the starting point of a continuous professional development scheme; indeed we considered continuing re-training as important as initial training. In fact, despite a widespread lack of ongoing training or other forms of continuing professional development, several studies have found the positive association between training and maintaining good standards of practices. (58–61). This is shown to be linked: (i) to refresh of acquired skills and knowledge and (ii) to motivation. For instance, Curtale et al. suggest that even few days additional training/supervision, if regularly provided, results in improved quality of service (62). Furthermore, access to training and supervision seems to be associated with non-monetary incentive sustaining CHWs motivation and engagement (13,61). Moreover, a continuing training centered on CHWs seems to be an asset to the strategy of continuous professional development. Puchalski Ritchie et al. in their training course, established training contents on needs they identified through a prior qualitative survey performed by CHWs (63). These recommendations sustain the design we adopted.

Ongoing training, including various moments of evaluation and reflection, enable interventions to be adjusted in time to ensure proper implementation. Constant efforts throughout the project are essential to maintain quality while adapting to the inevitable changes in the context in which the intervention evolves.

## 8 Author Contributions

CC, JMI, GM, MD and AS participated in the conceptualization. CC and JMI developed the methodology. CC made formal analysis and drafted the manuscript. JMI, MD and AS reviewed initial draft. All the authors read, corrected, and approved the final manuscript.

## Data Availability

All data produced in the present study are available upon reasonable request to the authors

## 10 Acknowledgments

Authors would like to thank all CHWs who participated to the training and the evaluation, and who are currently make possible the implementation of Curema intervention in the field. Moreover, authors thank Surinamese MoH and particularly the NMEP for their support to CHWs’ training program in Paramaribo.

## Data Availability Statement

Data available under reasonable request to the corresponding author.

